# Leveraging transfer learning from Acute Lymphoblastic Leukemia (ALL) pretraining to enhance Acute Myeloid Leukemia (AML) prediction

**DOI:** 10.1101/2025.09.17.25336037

**Authors:** Ajay Duraiswamy, David Harris-Birtill

## Abstract

We overcome current limitations in Acute Myeloid Leukemia (AML) diagnosis by leveraging a transfer learning approach from Acute Lymphoblastic Leukemia (ALL) classification models, thus addressing the urgent need for more accurate and accessible AML diagnostic tools.

AML has poorer prognosis than ALL, with a 5-year relative survival rate of only 17–19% compared to ALL survival rates of up to 75%, making early and accurate detection of AML paramount. Current diagnostic methods, rely heavily on manual microscopic examination, and are often subjective, time-consuming, and can suffer from inter-observer variability. While machine learning has shown promise in cancer classification, its application to AML detection, particularly leveraging the potential of transfer learning from related cancers like Acute Lymphoblastic Leukemia (ALL), remains underexplored.

A comprehensive review of state-of-the-art advancements in acute lymphoblastic leukemia (ALL) and acute myeloid leukemia (AML) classification using deep learning algorithms is undertaken and key approaches are evaluated. The insights gained from this review inform the development of two novel machine learning pipelines designed to benchmark effectiveness of proposed transfer learning approaches. Five pre-trained models are fine-tuned using ALL training data (a novel approach in this context) to optimize their potential for AML classification.

The result was the development of a best-in-class (BIC) model that surpasses current state-of-the-art (SOTA) performance in AML classification, advancing the accuracy of machine learning (ML)-driven cancer diagnostics.

**Author summary:** Acute Myeloid Leukemia (AML) is an aggressive cancer with a poor prognosis. Early and accurate diagnosis is critical, but current methods are often subjective and time-consuming. We wanted to create a more accurate diagnostic tool by applying a technique called transfer learning from a similar cancer, Acute Lymphoblastic Leukemia (ALL).

Two machine learning pipelines were developed. The first trained five different models on a large AML dataset to establish a baseline. The second pipeline first trained these models on an ALL dataset to ”learn” from it before fine-tuning them on the AML data. Our experiments showed that the models that underwent transfer learning process consistently performed better than the models trained on AML data alone. The MobileNetV2 model, in particular, was the best-in-class, outperforming all other models and surpassing the best-reported metrics for AML classification in current literature.

Our research demonstrates that transfer learning can enable highly accurate AML diagnostic models. The best-in-class model could potentially be used as a AML diagnostic tool, helping clinicians make faster and more accurate diagnoses, improving patient outcomes.

## Introduction

Acute Lymphoblastic Leukemia (ALL) and Acute Myeloid Leukemia (AML) are aggressive forms of leukemia that develop in the bone marrow, characterized by the presence of ”blasts,” or immature white blood cells that have undergone leukemic transformation [1]. The presence of blasts in peripheral blood smears at levels greater than 5% is associated with high mortality and comorbidities [2–4]. Both cancers carry a poor prognosis if detection is delayed. AML has a 5-year relative survival rate of only 17–19% [5, 6], while ALL survival rates range from 15–75% depending on the patient’s age at diagnosis [7].

The widespread use of deep learning models for identifying, classifying, and segmenting cancers in slide images is facilitated by the availability of curated image repositories such as The Cancer Imaging Archive (TCIA) [8]. While deep learning models capable of classifying millions of images have been developed [9], when image classification is performed on smaller datasets with limited variance, issues such as overfitting, biases [10], and memorization [11] can arise. These challenges often necessitate other machine learning techniques, such as transfer learning.

Transfer learning involves leveraging the weights of a trained CNN, learned from one problem, to improve performance on a different but related problem [12]. In cancer imaging, transfer learning uses a pre-trained network to build domain specific transfer knowledge and transfer this from one domain to improve classification accuracy in another.

### Suitability of Transfer Learning for AML and ALL

The morphological similarities between lymphoblasts (from Acute Lymphoblastic Leukemia, ALL) and myeloblasts (from Acute Myeloid Leukemia, AML) are also a key factor in their selection for this transfer learning research. While ALL and AML originate from different lineages (lymphoid vs. myeloid) [13], their blast cells share overlapping morphological features (as shown in Figs 1 and 2), including:

1. High nuclear-to-cytoplasmic ratio [15],
2. Round-to-oval nuclei with nucleoli [14]
3. Presence of azurophilic granules [15], i.e. organelles found in white blood cells that contain enzymes and proteins that help fight microbes [16].
4. Immature, undifferentiated appearance [17].

**Fig 1.** Lymphoblast (ALL) Morphology: Small to medium Size; High Nuclear to cytoplasmic ratio [14]

**Fig 2.** Myeloblast (AML) Morphology: Medium size, High Nuclear to cytoplasmic ratio; Azurophilic/Cytoplasmic granules; Auer Rod [14]

However, they also exhibit distinct morphological differences. The presence of Auer rods (needle-like crystals) is characteristic of myeloblasts but not lymphoblasts [15].

Myeloblasts are typically larger (15–20 µm) compared to the small to medium-sized lymphoblasts (10–15 µm). Irregular cell contours and a deeper blue cytoplasm are also unique to myeloblasts [18].

This balance of shared and distinct features makes myeloblasts and lymphoblasts well-suited for transfer learning research.

To the best of our knowledge, the application of transfer learning to leverage knowledge gained from one cancer type (ALL) to improve the detection and classification of a related but more aggressive cancer type (AML) [6] is a novel approach. AML has a significantly lower 5-year survival rate (31.9%) compared to ALL (72.0%) [19, 20], making improved AML detection particularly important. This research focuses on exploring this gap in the literature.

### Machine Learning in Leukaemia Image Classification

Machine Learning (ML) models, particularly Convolutional Neural Networks (CNNs) have been used extensively in automated identification of Lymphoblasts (from Acute Lymphoblastic Leukaemia, ALL) and myeloblasts (from Acute Myeloid Leukaemia, AML) [21].

#### AML Imaging Research

Recent studies in AML classification show a diverse range of approaches, from supervised approaches with labelled datasets to semi-supervised approaches. Matek et al. [22] use the AML-Cytomorphology_LMU dataset to report intra-class precision and recall of 94% and an average accuracy of >90%. Building on this, Lu et al. [23] proposed a novel Masked Autoencoder (MAE) combined with active learning (MAE4AL) on the AML dataset [24], achieving a classification accuracy of 96.4% with only 20% of training data, highlighting the potential of semi-supervised strategies. In contrast, Hu et al. [25] integrated RegNetX-3.2gf with a Cost-Sensitive Loss Function (CS) and Squeeze-and-Excitation Networks (SE), attaining modest performance (68.2% precision, 63.7% recall, 65.1% F1-score), suggesting challenges in class-imbalanced AML datasets. Meanwhile, Dasariraju et al. [26] use random forest algorithm for inter-class classification of mature and immature leukocytes, achieving 93.0% detection accuracy and 93.5% classification accuracy across four immature subtypes.

#### Mixed Imaging Research

Abhishek et al. [27] combined a fine-tuned VGG16 with SVM on a mixed dataset of 1,250 ALL, AML, CLL, and CML images, achieving 84% accuracy, while Ahmed et al. [28] trained a CNN on a smaller dataset (6,321 images) of similar composition to attain 81.7% accuracy. In contrast, Dese et al. [29] reported exceptional metrics (97.7% accuracy, 97.9% sensitivity, 100% specificity) using Support Vector Machine (SVM) on a heterogenous dataset; however, their reliance on a limited training set (∼250 images) and omission of evaluation results raise concerns about overfitting. Meanwhile, Baig et al. [30] achieved 97.4% accuracy via ensemble techniques (ANN & SVM, AlexNet, SABE), though the complexity of their multi-model approach may limit scalability.

Park et al. [31] employed EfficientNet-V2 on a large-scale dataset of 42,386 single-cell images but reported comparatively lower metrics (accuracy: 87.8%, F1: 72.1%), likely due to class imbalance or morphological ambiguity across leukaemia subtypes. The range of results highlight the considerations when balancing dataset diversity, model complexity, and dataset sizes in multi-class leukaemia diagnostics.

#### ALL Imaging Research

Das et al. [32] leveraged transfer learning with an orthogonal softmax layer on the C_NMC_2019 ALL dataset [33], achieving an F1-score of 89.5% and precision of 91%. Similarly, Rahmani et al. [34] combined DenseNet-201 with a hybrid RF-GABACO algorithm, attaining 90.4% accuracy and 95.9% sensitivity, while Pan et al. [35] employed a neighborhood-correction algorithm (NCA) on the same dataset, reporting a weighted F1-score of 92.5% in preliminary testing and 91.04% in final evaluations.

Building on their prior work [32], Das et al. [36] applied MobileNetV2-ShuffleNet fusion to the ALL-IDB1/2 datasets [37], achieving near-perfect metrics (e.g., 99.1% accuracy, 100.0% sensitivity on ALL-IDB1; 98.5% across all metrics on ALL-IDB2). However, Lin et al. [38] caution that such high performance on small datasets (ALL-IDB1: 108 images; ALL-IDB2: 206 images) may reflect over-memorization or catastrophic overfitting, necessitating robust validation. Earlier work by Das et al. [39] using SqueezeNet on the same datasets reported lower but more conservative metrics (96.0% accuracy, 92.6% sensitivity). Saikia et al. [40] achieved 98.4% accuracy via WOA-SVM, highlighting performance improvements from optimising traditional models. Similar to studies on AML and mixed datasets, these studies, too, highlight the tradeoffs between model complexity, and dataset scale in ALL diagnostics, with traditional models typically showing better performance.

Current approaches to acute leukemia (ALL/AML) classification show a diverse range of results in predictive accuracy. A consistently superior methodology, with repeatable results was not evident from this review. Therefore, we aim to addresses this gap by evaluating a suite of pre-trained deep learning architectures, and assessing how their feature extraction accuracy can be improved, focussing on AML classification.

### 0.1 Machine Learning Approaches for Knowledge Transfer

Several techniques address the limitations imposed by small medical imaging datasets on machine learning, such as domain adaptation [41–43], few-shot learning [44–46], multi-task learning [47], and ensemble methods [48, 49].

#### Domain Adaptation techniques

bridge the gap between source and target domains by learning domain-invariant representations. Adversarial domain adaptation methods use adversarial training to align feature distributions between domains [41], while other techniques, such as Deep CORAL (CORrelation ALignment), minimize domain differences through correlation alignment [42, 43]. However, domain adaptation typically assumes that source and target domains share the same label space, which may not hold when transferring knowledge between different cancer types (ALL vs AML) with subtle but varying classification criteria.

#### Few-Shot Learning

use minimal examples to adapt to new tasks by learning meta-knowledge from related tasks. Finn et al. [44], and Snell at al. [45] have demonstrated Model-Agnostic Meta-Learning (MAML) and prototypical networks as viable approaches in medical imaging classification. However, few-shot learning requires a diverse set of related tasks during training, and the limited availability of multiple leukaemia subtypes with sufficient data makes this approach challenging to implement effectively.

#### Multi-Task Learning

uses shared representations to learn across multiple related tasks [47]. Joint training on ALL and AML classification could theoretically benefit AML classification task. However, this approach requires balanced datasets, which is not possible given the significant disparity in available ALL and AML dataset sizes, i.e. 10,661 images vs 18,354 images respectively.

#### Ensemble Methods

combine models trained on different domains to improve generalization [48, 49]. This technique focuses on combining predictions rather than explicitly transferring learned representations between domains [50]. Ensemble models also do not address the challenge of limited target domain data (AML) if all ensemble members are trained primarily on the same limited AML dataset.

Transfer Learning is selected as the primary approach because it can leverage ALL image data to improve AML classification performance, despite disparity in dataset sizes. Also,transfer learning accommodates differences in label spaces, diagnostic criteria between cancer types, limited classification sub-types, dataset sizes, overcoming limitations of domain adaptation, few-shot learning, multi-task learning and ensemble techniques. Also, transfer learning has been used extensively to show significant improvement is classification performance over other medical imaging applications techniques, as seen in Table 2.

**Table 1.** Summarised Literature Review on ALL/AML Classification using Machine/Deep Learning.

| Authors | Ref. | Dataset Type | Dataset Size | Method |
| --- | --- | --- | --- | --- |
| Lu et al | [23] | AML | 18,365 | Masked Autoencoder (MAE) and active learning (AL) - MAE4AL |
| Hu et al. | [25] | AML | 18,365 | RegNetX-3.2gf coupled with Cost-Sensitive Loss Function (CS) and Squeeze-and-Excitation Networks (SE) |
| Dasariraju et al | [26] | AML | 18,365 | Random Forest Algorithm |
| Rahmani et al. | [34] | ALL | 15,114 | DenseNet 201 & Random Forest-Genetic Algorithm-Binary Ant Colony Optimisation (RF-GABACO) |
| Pan et al. | [35] | ALL | 10,661 | Neighborhood-Correction Algorithm (NCA) |
| Das et al. | [36] | ALL | 314 | MobileNetV2 coupled with ShuffleNet |
| Das et al. | [32] | ALL | 314 | SqueezeNet |
| Saikia et al. | [40] | ALL | 108 | Whale Optimisation Algorithm-based Support Vector Machine (WAO-SVM) |
| Park et al. | [31] | AML, ALL | 42,386 | EfficientNet-V2 (B2) |
| Dase et. al | [32] | ALL, ASH | ~1000 | Orthogonal SoftMax Layer (OSL) |
| Abhishek et al. | [27] | ALL, AML, CLL, CML | 1,250 | VGG16 with LTCL fine tuned along with SVM |
| Ahmed et al. | [28] | ALL, AML. CLL, CML | 6,321 | CNN |
| Dese et al. | [29] | ALL, AML. CLL, CML | 200 | SVM |
| Baig et al. | [30] | ALL, AML, MM | 4,150 | ANN & SVM, CNN & SVM, Active Contours, Alexnet and SABE on CCA Fusion |

**Table 2.**
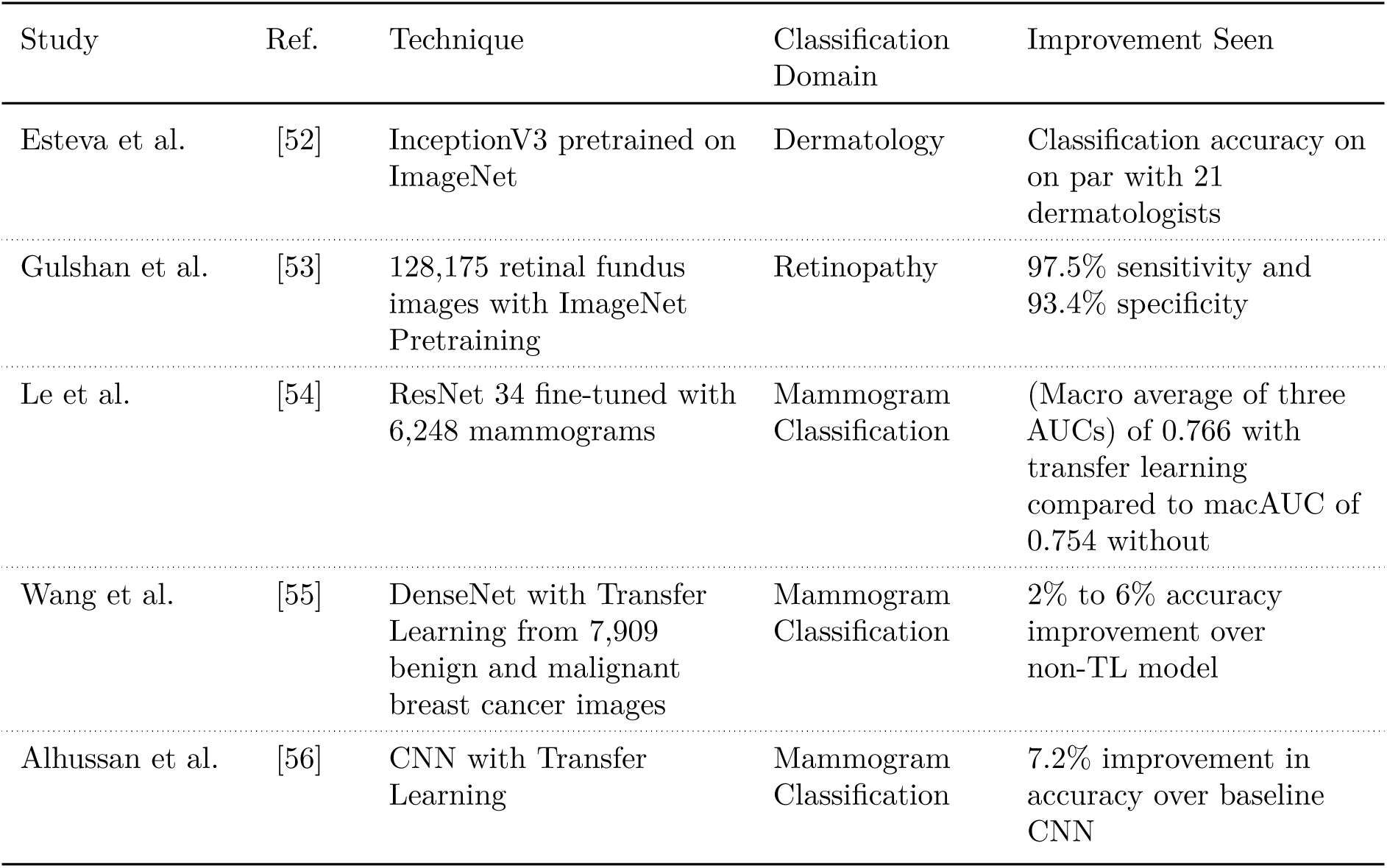
Summary of studies showing improvements in classification accuracy from implementing transfer learning.

#### Transfer Learning

Transfer learning involves the use of pre-trained networks to transfer learning from one domain to another related domain, overcoming the requirements for sufficiently large datasets [51], while mitigating over-fitting and memorisation issues risks that are seemingly common when DL networks are used with small datasets [11]

Two approaches are seen in literature, the first is the use of pretrained models, as feature extractors, directly on related image domains, without any domain-specific fine-tuning. The second approach involves fine-tuning a pre-trained model, with data from one domain to help improve classification performance on an analogous/related domain. Litjens et al. [51] show that fine-tuning pre-trained models yields superior classification performance in medical image analysis tasks relative to using pretrained models as direct feature extractors.

High classification accuracies are reported in literature when transfer learning methods are used. A summary of studies demonstrating performance improvements from utilising transfer learning are shown in Table 2

## Materials and methods

We selected five ImageNet pre-trained architectures: Vision Transformer [57], MobileNetV2 [58], DenseNet201 [59], EfficientNet-B0 [60], and ResNet101 [61]. Each model was first trained and tested on the AML dataset to establish baseline performance without transfer learning. Subsequently, models are pre-trained on the ALL dataset before fine-tuning on the AML dataset. These transfer-learned models are then evaluated on held-out AML test data to assess classification improvement. The best-performing model is benchmarked against state-of-the-art AML classification methods from literature. Following reproducibility guidelines highlighted by Fell et al. [62], we provide comprehensive details on model selection criteria, hyperparameter optimization procedures, and image preprocessing techniques to ensure transparency and reproducibility.

### ALL & AML Datasets

The data used in this research comes from two publicly available sources (under CC BY 3.0): ALL images from the Acute Lymphoblastic Leukemia (ALL) dataset, titled C-NMC-2019 [33] (referred to as the ”ALL Dataset”), and AML images from the Munich AML Morphology Dataset, titled AML-Cytomorphology-LMU [24] (referred to as the ”AML Dataset”). Following ethics approval obtained on 23/01/2025, formal data collection and analysis commenced using these identified datasets.

From the the ALL Dataset [33], we use 10,661 pre-classified, single-cell images. As shown in Table 3, 7,272 images are classified as leukemic lymphoblasts (ALL), while the remaining images depict non-cancerous cells. These 7,272 ALL images are split into training, validation, and testing (held-out) sets in a 74:22:4 ratio, respectively. The ALL Dataset images are in bitmap format with a resolution of 450×450 pixels and are pre-segmented, meaning non-cell areas are black.

**Table 3.** Overview of the ALL & AML Datasets used in this Research.

|  | Total | Train<br>74% |  | Validate<br>22% |  | Test<br>4% |  |
| --- | --- | --- | --- | --- | --- | --- | --- |
|  |  | Cancer | No Cancer | Cancer | No Cancer | Cancer | No Cancer |
| ALL | 10,661 | 5,351 | 2,494 | 1,601 | 746 | 320 | 149 |
| AML | 18,354 | 2,412 | 9,795 | 1,033 | 4,197 | 181 | 736 |

The AML Dataset consists of expert-labeled single-cell images from blood smears of 100 patients diagnosed with AML at Munich University Hospital between 2014 and 2017. It also includes images from 100 patients without any indications of blood-cell malignancy (control group). These images are in TIFF format with a resolution of 400×400 pixels.

Analysis of class balance, shown in Table 4 reveals that 38% of the total images (10,989) show cancer, while 62% (18,117) are not. This class imbalance requires specific mitigation strategies during model evaluation, which are discussed further.

**Table 4.**
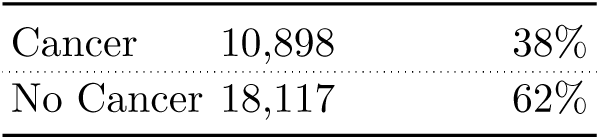
Class Balance in ALL & AML Datasets used in this Research.

The AML dataset contains blood smear images categorized into 15 morphological classes (see Table 5). A key objective of this research is to develop a state-of-the-art (SOTA) AML classifier that can assist in cancer detection (rather than performing intra-class morphological classification). Therefore, the classification task is framed as a binary problem: each of the 15 morphological classes is further categorized as either ”cancer” or ”non-cancer,” based on whether the presence of that class indicates an abnormality in blood smears.

**Table 5.** 2 Class Classification of Cell Type in AML Dataset.

| Acronym | Morphology | Type | Images |
| --- | --- | --- | --- |
| BAS | Basophil | Normal | 79 |
| EBO | Erythroblast | Cancer | 78 |
| EOS | Eosinophil | Normal | 424 |
| KSC | Smudge cell | Ignored |  |
| LYA | Lymphocyte (atypical) | Cancer | 11 |
| LYT | Lymphocyte (typical) | Normal | 3,937 |
| MMZ | Metamyelocyte | Cancer | 15 |
| MOB | Monoblast | Cancer | 26 |
| MON | Monocyte | Normal | 1,789 |
| MYB | Myelocyte | Cancer | 42 |
| MYO | Myeloblast | Cancer | 3,268 |
| NGB | Neutrophil (band) | Cancer | 109 |
| NGS | Neutrophil (segmented) | Normal | 8,484 |
| PMB | Promyelocyte (bilobed) | Cancer | 18 |
| PMO | Promyelocyte | Cancer | 70 |

**Table 6.** Rationale for Binary Classification of Cell Types in the AML Dataset.

| Cell Type | Description/Relevance to AML | Classification | Reference |
| --- | --- | --- | --- |
| Basophil | Typically indicates allergic reactions; rarely associated with chronic myeloid leukaemia | Non-Cancer | [63] |
| Erythroblast | Presence in peripheral blood suggests leukaemia | Cancer | [64] |
| Eosinophil | Usually linked to allergies/parasites; rare association with lymphomas | Non-Cancer | [65] |
| Lymphocyte (atypical) | May indicate lymphoma or Chronic Lymphocytic Leukaemia (CLL) | Cancer | [66] |
| Lymphocyte (typical) | Normal cells | Non-Cancer |  |
| Metamyelocyte | Immature forms in blood suggest AML | Cancer | [67] |
| Monoblast | Hallmark of AML | Cancer | [68] |
| Monocyte | Typically 2% to 8% of normal total white blood cell count | Non-Cancer | [69] |
| Myelocyte | Marker for AML | Cancer | [67] |
| Myeloblast | Definitive marker of Acute Myeloid Leukaemia (AML) | Cancer | [67] |
| Neutrophil (band) | Increased presence (bandemia) linked to AML | Cancer | [70] |
| Neutrophil (segmented) | Mature cells; non-specific for cancer | Non-Cancer |  |
| Promyelocyte (bilobed) | Characteristic of AML subtypes | Cancer | [67] |
| Promyelocyte | Key indicator of AML | Cancer | [71] |

**Table 7.**
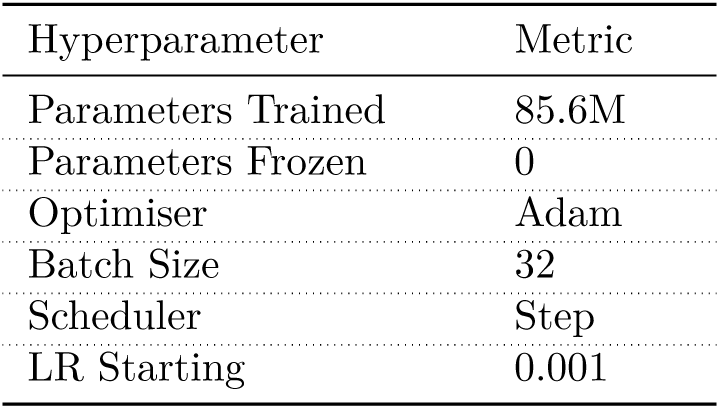
Vision Transformer Model Setup.

**Table 8.** EffcientNet V2 Model Setup.

| Hyperparameter | Metric |
| --- | --- |
| Parameters Trained | 2.2M |
| Parameters Frozen | 0 |
| Optimiser | Adam |
| Batch Size | 32 |
| Scheduler | Step |
| LR Starting | 0.001 |

**Table 9.**
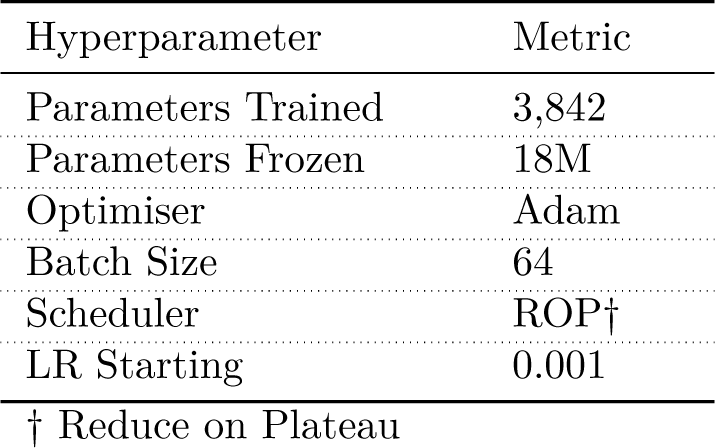
DenseNet201 Model Setup.

**Table 10.**
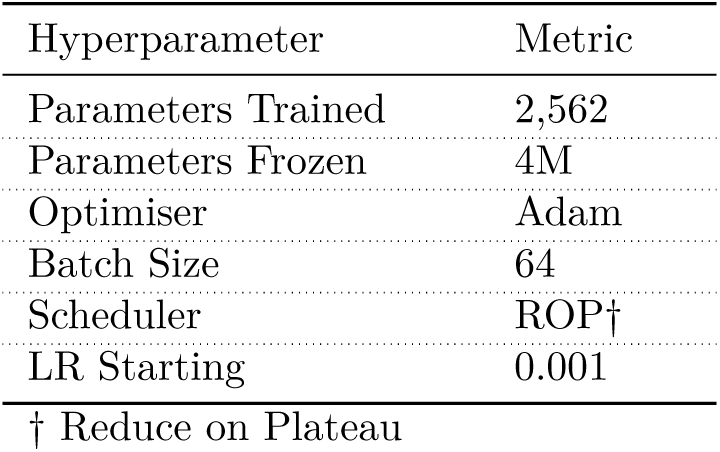
EfficentNet B0 Model Setup.

**Table 11.**
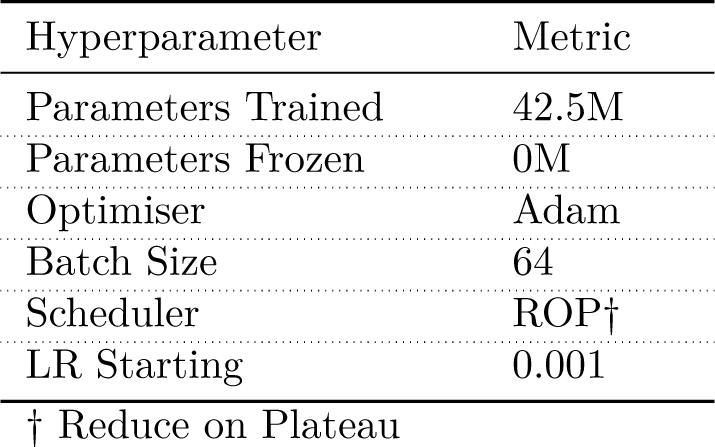
ResNet101 Model Setup.

| Hyperparameter | Metric |
| --- | --- |
| Parameters Trained | 42.5M |
| Parameters Frozen | 0M |
| Optimiser | Adam |
| Batch Size | 64 |
| Scheduler | ROP† |
| LR Starting | 0.001 |

† Reduce on Plateau

**Table 12.** AML Model Setup during Training phase.

| Model | Parameters<br>Trained | Parameters<br>Frozen | Opti-<br>miser | Batch<br>Size | Sched-<br>uler | Best<br>Epoch | Best<br>Epoch<br>LR | Training<br>Time<br>(s)** |
| --- | --- | --- | --- | --- | --- | --- | --- | --- |
| Vision Trfmr.† | 85.6M | 0 | Adam | 32 | Step | 16 | 1e-6 | 5887.5 |
| MobileNet V2 | 2.2M | 0 | Adam | 32 | Step | 50 | 0 | 1273.3 |
| DenseNet201 | 3,842 | 18M | Adam | 64 | ROP† | 23 | 5e-4 | 1682.6 |
| EfficientNetB0 | 2,562 | 4M | Adam | 64 | ROP† | 42 | 1e-3 | 1037.5 |
| ResNet 101 | 42.5M | 0 | Adam | 64 | ROP† | 37 | 6.3e-5 | 3540.1 |

† ROP: Reduce On Plateau ‡ Vision Tranformer uses vit\_base\_patch16\_224

**Table 13.** AML Model Performance on Validation Data.

| Model | Precision | Recall | F1 Score | Accuracy | Bal.<br>Accuracy |
| --- | --- | --- | --- | --- | --- |
| Vision T/fmr. | 92.1% | 90.3% | 91.2% | 94.5% | 90.3% |
| MobileNet V2 | 95.0% | 95.8% | 95.4% | 97.0% | 95.8% |
| DenseNet201 | 89.4% | 86.2% | 87.7% | 92.5% | 86.2% |
| EfficientNetB0 | 88.0% | 80.9% | 83.8% | 90.6% | 80.9% |
| ResNet 101 | 95.1% | 95.6% | 95.3% | 97.0% | 95.6% |

The details of how this binary classification approach was implemented is detailed in Table 6. Examples of images in each class is see in Fig 3 and Fig 4

**Fig 3.** Slide Image showing cancer indicating morphological variations in the AML Dataset (AML)

**Fig 4.** Slide Image showing non-cancer morphological variations in the AML Dataset (AML)

### Data Splits

Both the AML and ALL datasets were split into training, validation, and testing sets as shown in Table 3 and Fig 5.

**Fig 5.** Percentage and Number of Images across Dataset Splits

**Fig 6.** Image Pre-processing for ViT Model

**Fig 7.** Image Pre-processing for MobileNetV2 Model

**Fig 8.** Image Pre-processing for DenseNet201 Model

For the ALL dataset, 74% of the images (comprising 5,351 cancerous and 2,494 non-cancerous images) is allocated to the training set. The validation set consists of 22% of the data (1,601 cancerous and 746 non-cancerous images). The remaining 4% (320 cancerous and 149 non-cancerous images) formed the held-out test set. This same split ratio was applied to the AML dataset.

The specific data splits of 74%, 22%, and 4% for training, validation, and testing respectively is adopted for the following reasons:

1. **Comparability:** The ALL dataset, C-NMC-2019 [33], is made available publicly with this exact split ratio. By maintaining these ratios, the classification performance becomes comparable to other studies in literature that utilise this dataset.
2. **Transfer Learning Performance:** To enable transfer learning between the two datasets, the AML-Cytomorphology-LMU dataset [24] was split using ratios matching the predefined splits of the ALL dataset, the source dataset for transfer learning. Soekhoe et al. [72] show that the size of the target dataset impacts transfer learning performance, with comparable dataset sizes showing better transfer learning performance.

The resulting AML training set contains 9,795 cancerous and 2,412 non-cancerous images; the validation set has 4,917 cancerous and 1,033 non-cancerous images; and the held-out test set comprises 736 cancerous and 181 non-cancerous images.

### Model Selection Criteria

Five pre-trained models—Vision Transformer (ViT), MobileNetV2, DenseNet201, EfficientNetB0, and ResNet101—are selected. ViT excels at capturing global contextual relationships using self-attention [57], which is crucial for detecting subtle morphological anomalies in blood cells. MobileNetV2, with its lightweight depth-wise separable convolutions, reduces the risk of overfitting [58], a common issue with small datasets. DenseNet201 [59] enhances feature reuse via dense connectivity, enabling robust hierarchical feature extraction from complex cell structures. EfficientNet [60] optimises accuracy-efficiency trade-offs through compound scaling, balancing depth, width, and resolution-offering ideal flexibility. ResNet101 [61] addresses gradient degradation in deep networks via residual learning, enabling extraction of morphological features from small AML images.

ach model was sourced from the TIMM (TorchImageModels) library, [73], already pre-trained on large-scale datasets such as ImageNet-21k or ImageNet-1k. For this specific binary classification task, each model’s default 1,000-class classification head was replaced with a new 2-class classifier. First, the models were trained on the Acute Lymphoblastic Leukemia (ALL) dataset to acquire knowledge of ALL cancer cell features. They were then fine-tuned on the Acute Myeloid Leukemia (AML) dataset for the final classification task.

Their ImageNet pre-training provides a robust feature initialization, which has been shown to be beneficial in medical transfer learning [74]. This selection of models allows for a comprehensive evaluation of AML classification capabilities.

### Image Pre-Processing & Hyperparameter Selection

#### Vision Transformer (ViT)

The Vision Transformer (ViT) model, sourced from the TIMM (TorchImageModels) library [73], is pre-trained on ImageNet-21k (containing 14 million images and 21,000 classes) [57] and subsequently fine-tuned on ImageNet-1k. Since AML classification in this research is a binary task, the default model, originally designed for 1,000 classes, is modified by replacing the classification head with a 2-class classifier.

1. **(a)** Example Erythroblast from AML Dataset **(b)** Processed Image after transformations

#### Rationale

1. **Image Resizing:** The shorter edge of each image is resized to 256 pixels to standardise input sizes, which includes upscaling smaller images [75].
2. **Image Cropping:** A 224x224 pixel crop is used, as this is the standard input size for ImageNet pre-trained models [76].
3. **Tensors transformation** The raw pixel values are converted into a tensor format suitable for the ViT model. The pixel values are also normalized from the range [0, 255] to [0, 1] to ensure features are on a similar scale [77].
4. **Input Normalisation:** Since the pre-trained model weights are trained on ImageNet data (which has RGB channels with a mean of [0.485, 0.456, 0.406] and a standard deviation of [0.229, 0.224, 0.225] [78]), the input images are normalized to this same distribution. This helps prevent distribution shift and promotes faster convergence.

### Hyperparameters Selection

#### Rationale

1. **Full Fine-tuning:**In this ViT implementation, all 85.6 million parameters are trained (i.e., no layers are frozen). Freezing layers is generally less effective in ViTs compared to CNNs. Dosovitskiy et al. [57] suggest that ViT’s self-attention mechanism allows all layers to contribute to global feature integration. Furthermore, Chen et al. [79] find that full fine-tuning often yields better performance than partial freezing, even with limited data, as it allows the model to adapt more effectively to the target task.
2. **Linear LR Optimisation:** Smith [80] advocates for a linearly decreasing learning rate schedule. Other hyperparameters, such as the Adam optimiser, a starting learning rate of 0.001, and a batch size of 32, are commonly used for ViT implementations and are recommended in the literature, including [81].

#### MobileNetV2

The MobileNetV2 model provided by the TIMM (TorchImageModels) library [73] is pre-trained on ImageNet-1k (also referred to as ILSVRC 2012, a collection of 1.3 million images and 1,000 classes) [58]. Since AML classification in this research is a binary task, the default model, originally designed for 1,000 classes, is modified by replacing the classification head with a 2-class classifier.

**(a)** Example Erythroblast from AML Dataset **(b)** Processed Image after transformations,

### Denormalised & Adjusted

#### Rationale

1. **Image Resizing:** The shorter edge of each image is resized to 256 pixels to standardise input sizes, which includes upscaling smaller images [75].
2. **Image Cropping:** A 224x224 pixel crop is used, as this is the standard input size for ImageNet pre-trained models [76].
3. **Random Horizontal and Vertical Flips:** Images are randomly flipped horizontally and vertically with a given probability. This augmentation is used because left-right symmetry is not a distinguishing feature of cell images [82].
4. **Random Rotations:** Images are randomly rotated by ±15°. This is included because cell images may not be perfectly aligned [82].
5. **Tensor Transformation and Normalization:** Raw pixel values are converted into a tensor format suitable for the MobileNetV2 model. The pixel values are also normalized from the range [0, 255] to [0, 1] to ensure features are on a similar scale [77].
6. **Input standardisation:** Input features are standardised. This helps ensure that the pre-trained model weights (optimised on ImageNet data) can be effectively used for the target task (AML classification) without requiring retraining of the lower layers [83].

### Hyperparameters Selection

#### Rationale

1. **Full Fine-tuning:** In this MobileNets implementation, all 2.2 million parameters are trained. Similar to ViTs, full fine-tuning generally yields better performance than freezing layers, as it allows the model to adapt its inverted residuals, bottlenecks, and batch normalization layers during training [58].
2. **Linear LR Optimisation:** Smith [80] recommend a linearly decreasing approach to optimising learning rates. Other hyperparameters, such as the Adam optimiser, a starting learning rate of 0.001, and a batch size of 32, are commonly used for effective MobileNetV2 implementations and are recommended in the literature, including [84].

#### DenseNet201

The DenseNet201 model provided by the TIMM (TorchImageModels) library [73] is pre-trained on ImageNet-1k (also referred to as ILSVRC 2012, a collection of 1.3 million images and 1,000 classes) [59]. As modified in the previous instances, the default DenseNet201 model, originally designed for 1,000 classes, is modified by replacing the classification head with a 2-class classifier.

**(a)** Example Erythroblast from AML Dataset **(b)** Processed Image after transformations,

### Denormalised & Adjusted

#### Rationale

1. Rationale for Steps 1-5 are as stated in the above sections covering Vision Transformer & MobileNetV2.
2. **ColorJitter:** mitigates the potential impact of varying image capture settings on the DenseNet’s multiple transition layers, by randomly adjusting image brightness, contrast, and saturation within specified ranges—enabling more robust feature representations. [85].
3. **RandomErasing:** randomly sets rectangular regions of the image to arbitrary values, and encourages DenseNet to learn generalized feature representations, preventing over-reliance on specific image regions. DenseNets (dense) connections receives input from preceding layers. While this promotes feature reuse and strong performance, it can lead to overfitting if it learns heavily from a small image region [86].

### Hyperparameters Selection

#### Rationale

1. **Partial Fine-tuning:** In this DenseNet201 implementation, only the top 3,841 parameters of the custom classifier head are frozen, Mahapatra [87] find spatial inductive bias (translation invariance, locality) in Densenets is preserved when freezing early layers in DenseNet implementations.
2. **Discrete LR Optimisation:** Jastrzębski [88] suggest using discrete LR jumps, such as the Reducing LR on Plateau (ROP) strategy used in this research. The Adam optimiser and a starting learning rate of 0.001 were chosen as they are commonly recommended for DenseNet implementations in medical image classification [89].

#### EfficentNet B0

The EfficientNet-B0 model provided by the TIMM (TorchImageModels) library [73] is pre-trained on ImageNet-1k (also referred to as ILSVRC 2012, a collection of 1.3 million images and 1,000 classes) [60]. As modified in the previous instances, the default EfficientNet B0 model, originally designed for 1,000 classes, is modified by replacing the classification head with a 2-class classifier.

#### Image Preprocessing Steps

The pre-processing steps applied to the EfficientNet model is the same as described in the preceding sections for the other model implementations.

### Hyperparameters Selection

#### Rationale

1. **Partial Fine-tuning:** In this EfficientNet implementation, only the 2,562 parameters in the custom 2-channel classification head are trained; the remaining 4 million parameters are frozen. Tan and Le [60] recommend freezing the lower layers in EfficientNet, citing that the model relies on hierarchical feature extraction. The early layers, which capture low-level features like edges and textures, contribute to classification accuracy even when the model is fine-tuned on a specific task.
2. **Discrete LR Optimisation:** Jastrzębski [88] suggest using discrete LR jumps, such as the Reducing LR on Plateau (ROP) strategy used in this research. The choice of other hyperparameters, such as Adam for optimiser, starting learning rate of 0.001, are standard implementation approaches for Efficientnet, seen in literature, including [90].

#### ResNet101

The ResNet101 model provided by the TIMM (TorchImageModels) library [73] is pre-trained on ImageNet-1k (also referred to as ILSVRC 2012, a collection of 1.3 million images and 1,000 classes) [61]. As modified in the previous instances, the default ResNet101 model, originally designed for 1,000 classes, is modified by replacing the classification head with a 2-class classifier.

#### Image Preprocessing Steps

The pre-processing steps applied to the ResNet101 model is the same as described in the preceding sections for the other model implementations.

### Hyperparameters Selection

#### Rationale

1. **Full Fine-tuning:** In this ResNet101 implementation, all 42.5 million parameters are involved in the fine-tuning process. He et al. [61] argue that Residual Networks achieve stable training due to the leveraging short paths during training. These connections allow gradients to bypass layers, mitigating the risks of vanishing or exploding gradients. Furthermore, Veit et al. [91] suggest that ResNets behave like ensembles of shallower networks, exhibiting stable training and gradient flow, which further supports the decision to perform full-network training in this case.
2. **Discrete LR Optimisation:** Jastrzębski [88] suggest using discrete LR jumps, such as the Reducing LR on Plateau (ROP) strategy used in this research. The choice of other hyperparameters, such as Adam for optimiser, starting learning rate of 0.001, are all seen in standard implementation approaches for ResNet, as seen in [92].

#### F1 Score Primacy

F1 score is a harmonic mean of precision and recall, seen in Eq.1, and therefore penalises extreme values of either [93], and provides a balanced view of classification accuracy. It is generally considered a better metric than accuracy alone for evaluating classification algorithms with imbalanced datasets [94]. As this research uses an imbalanced dataset, F1 score is used as the primary criterion when selecting best performing models during evaluation process.

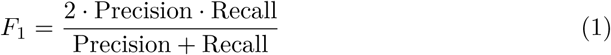

#### Model Training, Validation & Test Pipeline

As shown in Fig 9, the first step in the pipeline involves preprocessing the ALL and AML images from their respective datasets. The specific preprocessing steps vary depending on the deep learning model and are detailed in the preceding sections.

**Fig 9.** Methodology Showing Approach to Train-Validate-Test for Standalone and Transfer Learning Pipeline

#### AML Pipeline

Each deep learning model was trained and validated on the AML dataset using an AWS ml.g5.8xlarge cloud instance. The model-specific hyperparameter settings are as described in the previous section. Upon completion of each training job, the model’s performance metrics (precision, recall, F1 score, accuracy, and balanced accuracy) are recorded. The computation time was measured using the Python TicToC class and is included in the results presented in the sections below.

#### AML Standalone Evaluation

Following training, the models were evaluated on the held-out AML test dataset to assess their classification performance. Five key metrics were used to evaluate each model for AML classification performance.

### Transfer Learning Pipeline

#### ALL Initialisation

Each deep learning model was trained and validated on the ALL dataset using an AWS ml.g5.8xlarge cloud instance. The model-specific hyperparameter settings are as described in the previous section. After each training job, the model’s performance metrics (precision, recall, F1 score, accuracy, and balanced accuracy) is recorded.

#### AML Re-training

Following training, the weights from the epoch that yielded the best F1 score from ALL training were are to initialize the corresponding transfer learning model before retraining/ fine-tuning on the AML dataset (using an AWS ml.g5.8xlarge cloud instance).

After each training job, the model’s performance metrics (precision, recall, F1 score, accuracy, and balanced accuracy) is recorded.

#### Transfer Learning Evaluation

Finally, the five transfer-learned models are evaluated on the held-out AML test dataset to assess their AML classification performance. The same five key metrics were used to identify the best-in-class (BIC) transfer-learned model.

Following the recommendations of Muntean et al. [94] for evaluating classification algorithms on imbalanced datasets, the F1 score is used as the primary criterion for selecting the best-in-class (BIC) model, when similar metrics are seen across two or more models.

#### Benchmarking with SOTA on AML Classification

Once the best-in-class (BIC) model (i.e., the model exhibiting the highest AML classification performance) is identified, its performance is compared against state-of-the-art (SOTA) AML classification results reported in the literature to benchmark the proposed model.

#### Model & Code Availability

The code used in this research is available online at this Github link, and model weights are available at Zenodo [DOI:10.5281/zenodo.15388608] [95], and at Hugging Face storage repository to facilitate reproducibility of the presented results and ensure the validity of proposed models [62]

#### Ethics Approval

This research utilises two fully anonymised datasets from The Cancer Imaging Archive (TCIA) [8], analysed subsequent to ethics approval obtained on 23/01/2025. These datasets are - Acute Lymphoblastic Leukemia (ALL) dataset, titled, C-NMC-2019 [33] (referred to as the “ALL Dataset” in this research) consisting of 10,661 pre-classified, single-cell images, and the Munich AML Morphology Dataset, titled, AML-Cytomorphology-LMU [24] (referred to as the “AML Datset”), consisting of 18,354 single-cell images. From these single-cell images, no participants can be identified.

Clark et al [8] show that TCIA employs a rigorous de-identification process compliant with HIPAA’s Safe Harbor Method that anonymizes patient IDs and offsets dates using locally-retained mapping tables. All data transfers use encryption and are processed through RSNA’s Clinical Trial Processor software with scripts following the DICOM Attribute Confidentiality Profile standard. Upon receipt in a secured quarantine system, each image undergoes both visual inspection to ensure no PHI is burned into pixel data and automated analysis using Tag Sniffer software to verify complete removal of identifying information before public release.

Both datasets are available under the Creative Commons Attribution 3.0 Unported License, which allows for commercial, scientific, and educational use with proper attribution. Ethical approval for this study, including the use of aforementioned TCIA datasets, was also granted by the University of St Andrews, Computer Science Ethics Committee, Ref. No. 0040-CS-0040-29-2024.

## Results

### Standalone Classification Performance

#### Standalone AML Model

**AML Training & Validation** In the initial standalone experiment, five ImageNet pre-trained deep learning architectures—Vision Transformer, MobileNetV2, DenseNet201, EfficientNet, and ResNet101 are trained on the AML-Cytomorphology-LMU dataset [24] (referred to as the ”AML Dataset”). This dataset comprises 12,207 training images, including 2,412 cancerous and 9,795 non-cancerous images.

The performance metrics obtained during training each of the five models on the AML Dataset are presented in 13. The best metrics achieved in the AML standalone experiment are highlighted in purple indicate that both the MobileNetV2 and the ResNet101 model have learned the characteristics of AML morphologies comparably, and demonstrate strong overall performance (MobileNet V2 F1: 95.4% & ResNet101 F1: 95.3%) when classifying AML images, during training and validation.

Fig 10 and Fig 11 shows the P-R and loss plot of the MobileNetV2 and ResNet101 models (2 best models identified) during AML Standalone Model training.

**Fig 10.** P-R and loss Plot when Training Standalone MobileNetV2 (Best Model) with AML Dataset

**Fig 11.** P-R and loss Plot when Training Standalone Resnet101 (second-ranked model) with AML Dataset

**AML Model Evaluation** Subsequently, the best-performing model from each of the five AML-trained standalone models are evaluated on the held-out AML test dataset, which consists of 917 held-out (test) images (181 cancerous and 736 non-cancerous).

The ResNet101 model’s scores (F1: 95.4%; Precision: 95.0%; Recall: 95.9%; Accuracy: 97.1%; Bal. Accuracy: 95.9%), seen in Table 14 indicate that the ResNet101 model effectively learned the characteristics of AML morphologies and demonstrated the best overall performance, without pre-training (among the models compared in this experiment) in classifying AML cancer images, as shown in the confusion matrix for AML classification on the test data, see Fig 12.

**Fig 12.** Confusion Matrix/Heatmap for Evaluating Standalone Resnet101 on Held-out AML Test Dataset

**Table 14.** AML Model Performance on Test Data.

| Model | Precision | Recall | F1 Score | Accuracy | Bal.<br>Accuracy |
| --- | --- | --- | --- | --- | --- |
| Vision T/fmr. | 90.9% | 91.7% | 91.3% | 94.4% | 91.7% |
| MobileNet V2 | 94.7% | 95.8% | 95.2% | 97.0% | 95.8% |
| DenseNet201 | 89.5% | 88.6% | 89.0% | 93.1% | 88.6% |
| EfficientNetB0 | 89.6% | 82.7% | 85.5% | 91.6% | 82.7% |
| ResNet 101 | 95.0% | 95.9% | 95.4% | 97.1% | 95.9% |

#### Standalone ALL Model

**ALL Training & Validation** In the second standalone experiment, five ImageNet pre-trained deep learning architectures—Vision Transformer, MobileNetV2,

DenseNet201, EfficientNet, and ResNet101—were trained on the C-NMC-2019 dataset [33] (referred to as the ”ALL Dataset”). This dataset comprises 7,845 training images, consisting of 5,351 cancerous and 2,494 non-cancerous images.

Model-specific hyperparameters were selected and optimised, as detailed in the previous section. The best epoch, lowest learning rate, and training time achieved for each model are presented in Table 15. Consistent with the AML standalone training results, the EfficientNet B0 model exhibited the lowest training time (634.3 seconds) due to training only a small fraction of its parameters (2,562 out of 4 million).

**Table 15.** ALL Model Setup during Training phase.

| Model | Parameters<br>Trained | Parameters<br>Frozen | Opti-<br>miser | Batch<br>Size | Sched-<br>uler | Best<br>Epoch | Best<br>Epoch<br>LR | Training<br>Time<br>(s)** |
| --- | --- | --- | --- | --- | --- | --- | --- | --- |
| Vision Trfmr.† | 85.6M | 0 | Adam | 32 | Step | 15 | 1e-6 | 3690.6 |
| MobileNet V2 | 2.2M | 0 | Adam | 32 | Step | 21 | 0 | 815.4 |
| DenseNet201 | 3,842 | 18M | Adam | 64 | ROP† | 23 | 2.5e-4 | 1010.8 |
| EfficientNetB0 | 2,562 | 4M | Adam | 64 | ROP† | 42 | 1e-3 | 634.3 |
| ResNet 101 | 42.5M | 0 | Adam | 64 | ROP† | 37 | 6.3e-5 | 2226.6 |

† ROP: Reduce On Plateau ‡ Vision Tranformer uses vit\_base\_patch16\_224

The metrics from training the five models on ALL dataset is seen in Table 16. These best metrics achieved during the ALL standalone experiment are highlighted in purple indicate that the ResNet101 model has learned the characteristics of ALL morphologies comparably better, and demonstrates strong overall performance (F1=94.6%) in classifying images, during training and validation.

**Table 16.** ALL Model Performance on Validation Data.

| Model | Precision | Recall | F1 Score | Accuracy | Bal.<br>Accuracy |
| --- | --- | --- | --- | --- | --- |
| Vision T/fmr. | 85.0% | 81.0% | 82.5% | 85.6% | 81.0% |
| MobileNet V2 | 93.5% | 91.4% | 92.4% | 93.5% | 91.4% |
| DenseNet201 | 86.0% | 77.4% | 79.9% | 84.4% | 77.4% |
| EfficientNetB0 | 80.8% | 79.7% | 80.2% | 83.1% | 79.7% |
| ResNet 101 | 95.7% | 93.8% | 94.6% | 95.4% | 93.8% |

Fig 13 shows the P-R and loss plot of the ResNet101 model (best model identified) during ALL Standalone Model training.

**Fig 13.** P-R and loss Plot during Training Standalone ResNet101 with ALL Dataset

**ALL Model Evaluation** In the next step, the best-performing model from each of the five ALL-trained standalone models is evaluated on the held-out ALL test dataset, which consists of 469 unseen (test) images (320 cancerous and 149 non-cancerous).

ResNet101’s scores (F1: 95.3%; Precision: 95.9%; Recall: 94.7%; Accuracy: 96.0%; Bal. Accuracy: 94.7%), seen in Table 17 indicates that in this experiment too, the Resnet 101 model has learned the characteristics of ALL morphologies well and demonstrates best overall performance (among other models compared in this experiment) in classifying ALL cancer images, seen in the confusion matrix, in Fig 14.

**Fig 14.** Confusion Matrix/Heatmap from Evaluating Standalone Resnet101 on Held-out ALL Test Dataset

**Table 17.** ALL Model Performance on Test Data.

| Model | Precision | Recall | F1 Score | Accuracy | Bal. Accuracy |
| --- | --- | --- | --- | --- | --- |
| Vision T/fmr. | 81.6% | 81.5% | 81.5% | 84.0% | 81.5% |
| MobileNet V2 | 95.6% | 93.5% | 94.5% | 95.3% | 93.5% |
| DenseNet201 | 86.0% | 82.3% | 83.7% | 86.6% | 82.3% |
| EfficientNetB0 | 80.3% | 80.3% | 80.3% | 82.9% | 80.3% |
| ResNet 101 | 95.9% | 94.7% | 95.3% | 96.0% | 94.7% |

## Transfer Learning Classification Performance

### ALL **→** AML Transfer Learning Model

#### ALL **→** AML Training & Validation

The third phase of the experiment involves two sequential training steps to evaluate the benefits of transfer learning. Following the standalone ALL model training described in the previous section, each of the five ImageNet pre-trained deep learning architectures (Vision Transformer, MobileNetV2, DenseNet201, EfficientNet, and ResNet101), now initialized with the best weights from the ALL training, are subsequently fine-tuned on the AML dataset.

Model-specific hyperparameters are selected and optimised, as detailed in the previous section. The best epoch, lowest learning rate, and training time achieved for each model during this transfer learning phase are presented in Table 18.

**Table 18.** ALL → AML TL Model Setup during Training phase.

| Model | Parameters<br>Trained | Parameters<br>Frozen | Opti-<br>miser | Batch<br>Size | Sched-<br>uler | Best<br>Epoch | Best<br>Epoch<br>LR | Training<br>Time<br>(s)** |
| --- | --- | --- | --- | --- | --- | --- | --- | --- |
| Vision Trfmr.† | 85.6M | 0 | Adam | 32 | Step | 16 | 1e-6 | 5895.0 |
| MobileNet V2 | 2.2M | 0 | Adam | 32 | Step | 21 | 0 | 1330.6 |
| DenseNet201 | 3,842 | 18M | Adam | 64 | ROP† | 25 | 2.5e-4 | 1736.0 |
| EfficientNet | 2,562 | 4M | Adam | 64 | ROP† | 18 | 1e-3 | 1036.0 |
| ResNet 101 | 42.5M | 0 | Adam | 64 | ROP† | 44 | 3.1e-5 | 3547.8 |

† ROP: Reduce On Plateau ‡ Vision Transformer uses vit\_base\_patch16\_224

The metrics from training the transfer-learned models is seen in Table 19. These best metrics achieved during transfer-learning training is highlighted in purple indicate that the ALL-weighted - ResNet101 model has learned the characteristics of AML morphologies comparably better, and demonstrates strong overall performance (F1=95.5%) in classifying images, during AML training and validation.

**Table 19.** ALL → AML TL Model Performance on Validation Data.

| Model | Precision | Recall | F1 Score | Accuracy | Bal.<br>Accuracy |
| --- | --- | --- | --- | --- | --- |
| Vision T/fmr. | 92.4% | 91.6% | 92.0% | 95.0% | 91.6% |
| MobileNet V2 | 94.7% | 95.7% | 95.1% | 96.9% | 95.7% |
| DenseNet201 | 89.5% | 86.6% | 88.0% | 92.6% | 86.6% |
| EfficientNet | 88.1% | 83.0% | 85.2% | 91.2% | 83.0% |
| ResNet 101 | 95.2% | 95.9% | 95.5% | 97.2% | 95.9% |

Fig 15 shows the P-R and loss plot of the ResNet101 models (best model identified) during ALL → AML TL Model training.

**Fig 15.** P-R and loss Plot for Training MobileNetV2 ALL → AML TL Model

#### ALL **→** AML Transfer Learning Model Evaluation

As the final step in the experiment, each of the five ALL-to-AML transfer learning models are evaluated on the held-out AML test dataset, consisting of 917 held-out test images (181 cancerous and 736 non-cancerous), to assess the classification improvements achieved by the transfer learning approach compared to the standalone models.

These results indicate that in the transfer learning experiment, the MobileNetV2 and ResNet101 models demonstrated comparably strong performance in AML classification, with F1 scores of 96.2% and 96.1%, respectively.

The comparable metrics, seen in Table 20 suggest that both models effectively learned the characteristics of AML morphologies and exhibited strong overall performance (relative to the other models in this experiment) in classifying AML cancer images. While the MobileNetV2 transfer learning (TL) model outperformed the ResNet101 TL model in precision, F1 score, and accuracy, the ResNet101 TL model achieved higher scores in recall and balanced accuracy.

**Table 20.** ALL → AML TL ResNet101 Model Performance on Test Dataset.

| Model | Precision | Recall | F1 Score | Accuracy | Bal.<br>Accuracy |
| --- | --- | --- | --- | --- | --- |
| Vision T/fmr. | 93.5% | 93.0% | 93.3% | 95.8% | 93.0% |
| MobileNet V2 | 96.0% | 96.4% | 96.2% | 97.6% | 96.4% |
| DenseNet201 | 90.2% | 88.4% | 89.3% | 93.4% | 88.4% |
| EfficientNet | 88.2% | 84.4% | 86.1% | 91.6% | 84.4% |
| ResNet 101 | 95.6% | 96.6% | 96.1% | 97.5% | 96.6% |

**Table 21.** ALL → AML TL - Vision Transformer Model Performance on Test Dataset.

| Metrics \ Model | Vision Transformer |  |  |
| --- | --- | --- | --- |
|  | Standalone |  | Transfer Learning |
|  | ALL Only | AML Only | ALL → AML |
| Precision | 81.6% | 90.9% | 93.5% |
| Recall | 81.5% | 91.7% | 93.0% |
| F1 Score | 81.5% | 91.3% | 93.2% |
| Accuracy | 84.0% | 94.4% | 95.8% |
| Balanced Accuracy | 81.5% | 91.7% | 93.0% |

**Table 22.** ALL → AML TL - MobileNet V2 100 Model Performance on Test Dataset.

| Metrics \ Model | MobileNet V2 100 |  |  |
| --- | --- | --- | --- |
|  | Standalone |  | Transfer Learning |
|  | ALL Only | AML Only | ALL → AML |
| Precision | 95.6% | 94.7% | 96.0% |
| Recall | 93.5% | 95.8% | 96.4% |
| F1 Score | 94.5% | 95.2% | 96.2% |
| Accuracy | 95.3% | 97.0% | 97.6% |
| Balanced Accuracy | 93.5% | 95.8% | 96.4% |

**Table 23.** ALL → AML TL - DenseNet Model Performance on Test Dataset.

| Metrics \ Model | DenseNet 201 |  |  |
| --- | --- | --- | --- |
|  | Standalone |  | Transfer Learning |
| | ALL Only | AML Only | ALL → AML |
| Precision | 86.0% | 89.5% | 90.2% |
| Recall | 82.3% | 88.6% | 88.4% |
| F1 Score | 83.7% | 89.0% | 89.2% |
| Accuracy | 86.6% | 93.1% | 93.3% |
| Balanced Accuracy | 82.3% | 88.6% | 88.4% |

**Table 24.**
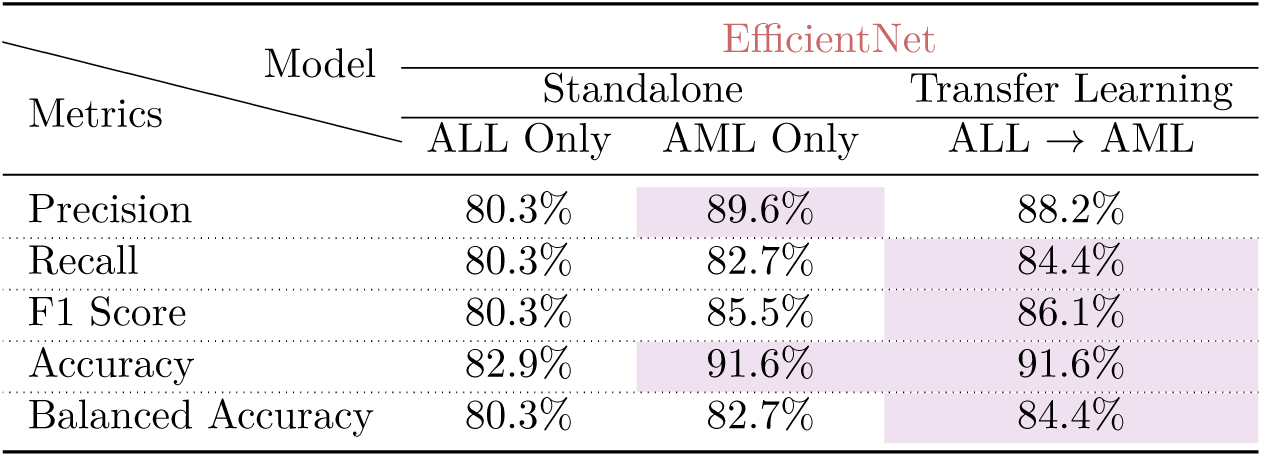
ALL → AML TL - EfficientNet Model Performance on Test Dataset.

**Table 25.**
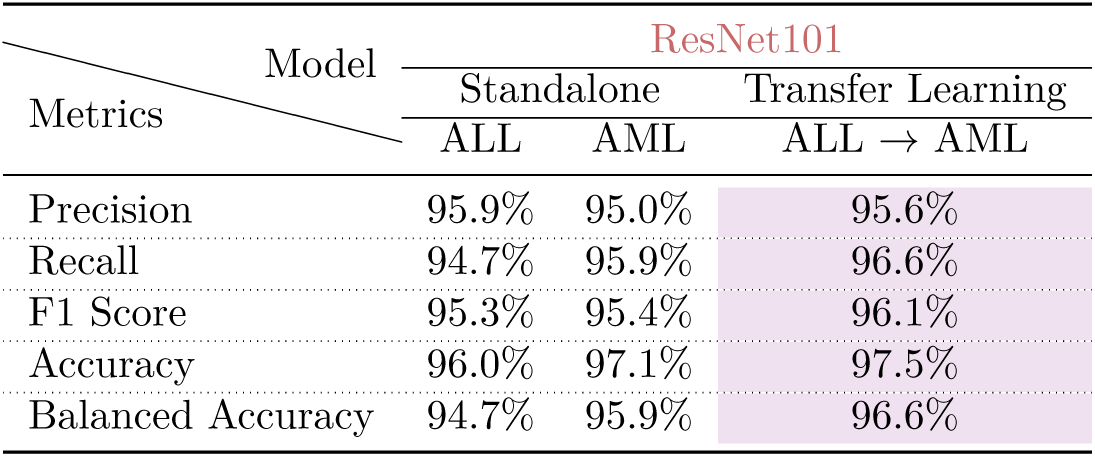
ALL → AML TL - ResNet101 Model Performance on Test Dataset.

| Metrics \ Model | ResNet101 |  |  |
| --- | --- | --- | --- |
|  | Standalone |  | Transfer Learning |
| | ALL | AML | ALL → AML |
| Precision | 95.9% | 95.0% | 95.6% |
| Recall | 94.7% | 95.9% | 96.6% |
| F1 Score | 95.3% | 95.4% | 96.1% |
| Accuracy | 96.0% | 97.1% | 97.5% |
| Balanced Accuracy | 94.7% | 95.9% | 96.6% |

**Table 26.** ALL → AML TL - Best-in-Class (BIC) Model Performance on Test Dataset.

| Metrics \ Model | ALL → AML TL Models Compared | | | | |
| --- | --- | --- | --- | --- | --- |
|  | ViT | MobileNet V2 | DenseNet 201 | EfficientNet | ResNet101 |
| Precision | 93.5% | 96.0% | 90.2% | 88.2% | 95.6% |
| Recall | 93.0% | 96.4% | 88.4% | 84.4% | 96.6% |
| F1 Score | 93.2% | 96.2% | 89.2% | 86.1% | 96.1% |
| Accuracy | 95.8% | 97.6% | 93.3% | 91.6% | 97.5% |
| Balanced Accuracy | 93.0% | 96.4% | 88.4% | 84.4% | 96.6% |

Because MobileNetV2 outperformed ResNet101 in three of the five evaluation metrics, and given the prioritization of the F1 score (as explained in the methods section), MobileNetV2 is considered to have demonstrated superior overall classification performance in the transfer learning experiment, emerging as the Best-in-Class (BIC) model.

The confusion matrix showing the (BIC) transfer-learned MobileNet V2’s classification performance on AML test data is seen in Fig 16.

**Fig 16.** Confusion Matrix/Heatmap for ALL → AML MobileNetV2 Transfer Learned Model on Held-out Test Dataset

#### Transfer Learning Improvements

The performance metrics of the transfer learning models are compared to those of their respective standalone ALL and AML models. Across all tested models, the transfer learning models AML showed improvements in F1 score compared to their standalone AML counterparts. In a significant number of cases, the transfer learning models also exhibited improved performance in the other four metrics: precision, recall, accuracy, and balanced accuracy. In the tables below, superior scores (compared between standalone AML and transfer-learned AML model) are shown in purple.

#### Benchmarking with SOTA on AML Classification

In the previous section, the performance metrics of the transfer learning models were compared with those of the respective standalone ALL and AML models to identify the Best-in-Class (BIC) model (using the F1 score as the primary discriminating factor).

This section benchmarks the BIC model against state-of-the-art (SOTA) AML classification results reported in current literature.

Table 27 below compares the metrics of the BIC transfer learning model (MobileNetV2) with those reported in the literature are enclosed in the Table 27 demonstrates that the proposed model’s metrics significantly exceed those of SOTA approaches reported in the literature for AML classification. This improvement is observed even when comparing results obtained using the same dataset (AML-Cytomorphology-LMU) [24]. A detailed legend is provided within Table 27.

**Table 27.**
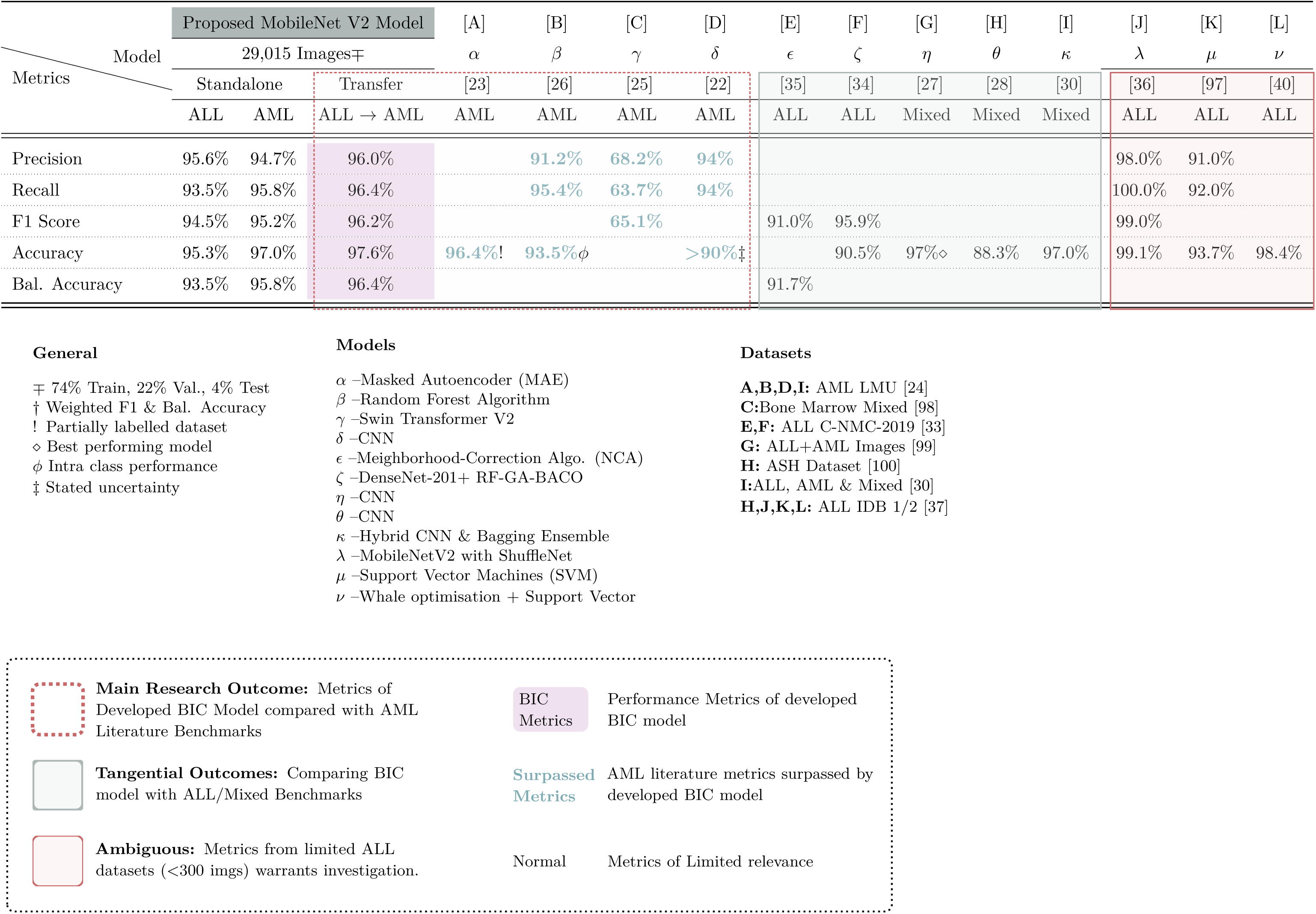
Comparison of Proposed Model Performance versus SOTA Reported in Literature.

## Discussion

To test the hypothesis that transfer learning (from ALL and ImageNet pre-training) can improve classification performance in AML image classification, with ALL pre-training, five ImageNet pre-trained DL architectures were selected: Vision Transformer, MobileNetV2, DenseNet201, EfficientNet, and ResNet101. Each model required unique hyperparameter tuning, including learning rate schedulers, starting learning rates, and the number of trainable parameters. As the objective is to identify a Best-in-Class (BIC) model, fine-tuning hyperparameters for individual model types does not affect this overall goal.

Two training pipelines were established, Fig 9). The first, the ”standalone” pipeline, trained each of the five models as standalone ALL and AML classifiers, respectively. This allowed for performance assessment without the benefit of transfer learning. In the second, the ”transfer learning” pipeline, each of the five models are first pre-trained on the ALL dataset and then fine-tuned on the AML dataset. These transfer-learned models were then evaluated on held-out AML test data to assess the impact of transfer learning on AML classification accuracy. Training time was recorded and reported as part of results, in accordance with best practice recommendations [96].

### AML Standalone Experiment

In the AML standalone experiment, where each of the five models was trained on the AML dataset, the MobileNetV2 model achieved better AML classification metrics on the training and validation sets. However, on the test data, the ResNet101 model outperformed the other AML-trained models. Arpit et al. [11] suggest that DL models learn patterns before memorizing noise, which could explain the differing performance on training/validation versus test data. Since evaluations are performed on (held-out) test data (which better reflects real-world performance), the standalone ResNet101 model emerged as the best-performing model overall, despite MobileNetV2’s superior performance on the validation set. The confusion matrix/heatmap (Fig. 12) highlights ResNet101’s strong AML classification performance, with 720 of 736 (97.8%) correctly predicted true negatives and 170 of 181 (93.9%) correctly predicted true positives on the held-out test dataset.

### ALL Standalone Experiment

The next experiment, the ALL standalone experiment, repeated the previous steps, but with each of the five models trained on the ALL training and validation sets. Hyperparameters, learning rate schedulers, starting learning rates, and the number of trainable parameters were again tuned for each model. Here, the ResNet101 model demonstrated superior ALL classification metrics on both the validation and test datasets. The confusion matrix (Fig. 14) shows this performance, with 136 of 149 (91.2%) correctly predicted true negatives and 314 of 320 (98.1%) correctly predicted true positives on the test data.

### ALL **→**AML Transfer Learning Experiment

The final experiment consisted of two sequential training steps. Following the ALL standalone training, each of the five ImageNet pre-trained architectures (Vision Transformer, MobileNetV2, DenseNet201, EfficientNet, and ResNet101), initialized with the best weights from the ALL training, was fine-tuned on the AML dataset. On the validation data, ResNet101 exhibited better classification metrics. However, on the test data, MobileNetV2 and ResNet101 showed comparable performance. While MobileNetV2 transfer-learned (TL) model outperformed ResNet101 TL model in precision, F1 score, and accuracy, ResNet101 TL model excelled in recall and balanced accuracy.

As established previously, the F1 score was the primary criterion for selecting the best-performing models due to its suitability for imbalanced datasets [94]. Based on the F1 score, the MobileNetV2 model was identified as the best-in-class (BIC) AML classification model, surpassing ResNet101 in precision (96.0%), F1 score (96.2%), and accuracy (97.6%).

The confusion matrix (Fig 16) on the test data reflects MobileNetV2’s strong classification performance on the held-out AML images (test dataset), with 724 of 736 (98.3%) correctly predicted true negatives and 171 of 181 (94.4%) correctly predicted true positives.

These results are also significant because they demonstrate superior performance with MobileNetV2,a model with only 2.2M parameters, compared to the denser models, e.g. ResNet101 (42.5M parameters), Vision Transformer (85.6M parameters). This contrasts with some studies suggesting that deeper networks typically achieve higher accuracy than models with fewer parameters [101, 102].

### Understanding MobileNetV2’s Results

This section explores the reasons for MobileNetV2’s class-leading performance compared to the other selected models.

### Lightweight Architecture and Reduced Overfitting

MobileNetV2 is the lightest model in the evaluated set, and among the most lightweight DL models available. It has 2.2 million trainable parameters, compared to 85.6 million for Vision Transformer, 18.09 million for DenseNet201, 4 million for EfficientNet, and 42.5 million for ResNet101 [57–61].

MobileNetV2’s design, prioritising efficiency with depthwise separable convolutions and inverted residuals [58], makes it less susceptible to overfitting on small medical datasets [103]. Larger models like ViTs or DenseNets may not be as well-suited to the detailed information contained within smaller blood cell images. Howard et al. [104] too show that smaller models like MobileNetV2 perform better on smaller datasets and are less prone to overfitting.

### Feature Reusability for Cell Morphology

Compared to ViTs, MobileNetV2’s inverted residual blocks efficiently capture hierarchical features—well-suited for leukaemia classification, which involves correctly identifying the thin envelope of pale blue cytoplasm surrounding the blast cell [4].

Further, Raghu et al. [105] show that ViTs learn differently than CNNs and can struggle with textures and local spatial features (crucial for this use case), often requiring more data than CNNs.

The automated scaling feature of EfficientNet [60] may have led to the prioritisation of unnecessary image features, potentially contributing to the poorer classification performance observed in these experiments.

### Data Efficiency

Dosovitskiy et al. [57] suggest that CNNs often perform better than ViTs on smaller datasets. While 20,052 images were used in training, the over-parameterization of DenseNet, ResNet, and ViT might have benefited from larger datasets for optimal results.

### Regularization

MobileNetV2’s inherent regularization (through bottleneck layers and fewer parameters) reduces the need for explicit regularization techniques like dropout [58]. Bottleneck layers omit the activation function, preserving information, avoiding issues such as exploding and vanishing gradients, which may have affected other models in this experiment.

### Class Imbalances

MobileNetV2’s lower complexity was better equipped to handle the 62:38 (No Cancer: Cancer) class imbalance in the dataset without requiring advanced sampling techniques, such as oversampling, suggested by Buda et al. [106].

In summary, the transfer-learned MobileNetV2 model exhibits best-in-class (BIC) AML classification performance, achieving precision, F1 score, and accuracy of 96.0%, 96.2%, and 97.6%, respectively. On the held-out test data, MobileNetV2 demonstrated strong classification performance, with 724 of 736 (98.3%) correctly predicted true negatives and 171 of 181 (94.4%) correctly predicted true positives, making it a promising candidate for a decision-support system in cancer diagnostics.

This section explored some of the reasons for MobileNetV2’s BIC performance. The next section discusses the performance of this best-in-class (BIC) model when benchmarked against state-of-the-art AML classification results reported in the literature.

### Benchmarking with SOTA AML Results

Lu et al. [23], using the same AML Cytomorphology LMU dataset [24] and a Masked Autoencoder with Active Learning (MAE), achieved 96.4% accuracy. While this is a strong result, the MobileNetV2 model proposed in this research surpasses it with 97.2% accuracy. Dasariraju et al. [26], also using the AML Cytomorphology LMU dataset, employed a Random Forest algorithm and achieved precision, recall, and accuracy of 91.2%, 95.4%, and 93.5%, respectively, on intra-class classification of AML cell subtypes. These metrics are lower than those achieved by the proposed BIC model (see Table 28). Hu et al. [25], using the larger MLL Helmholtz Fraunhofer dataset of 171,375 cell images representing various hematological diseases and a Swin Transformer V2, reported precision, recall, and F1 score of 68.2%, 63.7%, and 65.1%, respectively, on AML classification task. The proposed BIC model significantly outperforms these metrics.

**Table 28.** Best-in-Class MobileNetV2 Model Proposed for AML Classifcation.

|  |  |
| --- | --- |
| Precision | 96.0% |
| Recall | 96.4% |
| F1 Score | 96.2% |
| Accuracy | 97.6% |
| Bal. Accuracy | 96.4% |

Matek et al. [22] also used the AML Cytomorphology LMU dataset and reported an average precision and recall of 94% and an average accuracy of >90% for intra-class classification of AML cell subtypes. However, their reported precision varied widely, from 0.99 ± 0.00 for Neutrophils (segmented) to 0.07 ± 0.13 for Metamyelocytes, and their recall ranged from 0.96 ± 0.01 for Neutrophils to 0.07 ± 0.13 for Lymphocytes (atypical). The proposed BIC model exceeds the average metrics stated, although the focus here is not intra-class classification.

### Benchmarking with SOTA ALL Results Tangential Findings

While not an identified focus of this research, the performance of the standalone ALL ResNet101 model (see Table 29) is compared with some state-of-the-art ALL classification metrics reported in the literature to provide a broader context. Notably, the proposed ALL ResNet101 model achieves comparable or superior performance to some reported results, despite being trained without transfer learning or fine-tuning.

**Table 29.** Standalone ALL ResNet101 Model Proposed for ALL Classification Precision 95.9%.

|  |  |
| --- | --- |
| Precision | 95.9% |
| Recall | 94.7% |
| F1 Score | 95.3% |
| Accuracy | 96.0% |
| Balanced Accuracy | 94.7% |

Pan et al. [35], using the C-NMC-2019 dataset [33] and a Neighborhood Correction Algorithm, achieved an F1 score of 91.0% and a balanced accuracy of 91.7%. Both of these metrics are surpassed by the proposed standalone ALL ResNet101 model. Rahmani [34], also use the C-NMC-2019 dataset, and DenseNet-201 with RF-GA-BACO (Random Forest-Genetic Algorithm-Binary Ant Colony optimiszation) to achieve an accuracy of 90.5%, also outperformed by the developed ALL model.

It is important to note that some results (highlighted in red in the table) are not directly comparable because they use a much smaller dataset (100-300 images) called ALL-IDB1/2 [37]. Lin et al. [38] caution that ”over-memorization” and ”catastrophic overfitting” are prevalent on models trained using very small datasets. Given the limited size of the ALL-IDB1 (108 images) and ALL-IDB2 (206 images) datasets, the high metrics reported using these datasets may warrant further investigation to ensure they are not a result of overfitting.

Finally, the results reported under the ”mixed” data columns by [27, 28, 30] are not directly comparable as they use datasets containing a mix of leukaemia types (including Chronic Myeloid Leukaemia (CML) and Chronic Lymphoblastic Leukaemia (CLL)). These results are included for informational purposes only.

### Limitations

This research has some limitations, primarily the limited number of datasets used for the leukaemia conditions explored. Another limitation is the number of models evaluated, particularly smaller models that leverage the compact feature representation of the best-in-class MobileNetV2 model (such as SqueezeNet with 1.1 million parameters), could be explored. Sandler et al. [58] suggest that model compactness is crucial for feature learning when dealing with small object sizes, such as cell images.

### Future Work

Future work could also involve expanding the scope to include other related leukaemia conditions, such as Chronic Lymphoblastic Leukaemia (CLL) and Chronic Myeloid Leukaemia (CML), to investigate whether incorporating these cell morphologies contributes to enhanced transfer learning for AML classification. Conversely, the transfer learning direction could be reversed, exploring the impact of AML pre-training on subsequent ALL classification performance.

## Conclusion

We investigated whether learning from one type of cancer (ALL) could improve the detection performance for a related cancer type (AML).

Training data, consisting of 29,015 images from two publicly available ALL and AML datasets, was obtained from The Cancer Imaging Archive (TCIA). Five ImageNet pre-trained deep learning architectures—Vision Transformer, MobileNetV2, DenseNet201, EfficientNet, and ResNet101, were selected based on their complementary strengths in medical image classification. Model-specific hyperparameters and tuning strategies (schedulers, optimisers) were implemented based on best practices reported in the literature. Customized preprocessing steps were applied to the training images to meet each model’s specific requirements.

Two distinct deep learning pipelines were developed. The first pipeline assessed the models’ ability to classify AML images when trained solely on AML data (without transfer learning). The second pipeline utilised weights from ALL training to enhance AML classification. This process was repeated for each of the five selected models, and metrics were calculated to assess AML classification performance on an held-out test dataset.

Across all tested models, the transfer learning approach resulted in improved F1 scores compared to the standalone models. In many cases, improvements were also observed in the other four metrics: precision, recall, accuracy, and balanced accuracy.

The best-in-class model (the transfer-learned MobileNetV2) was compared against state-of-the-art AML classification results reported in the literature to benchmark its performance. The proposed BIC model surpassed previously reported AML classification metrics, including studies that use the same AML training data.

Some secondary outcomes were also observed, i.e. the classification performance of the standalone ALL ResNet101 model was compared with state-of-the-art ALL classification metrics. Notably, the standalone ALL ResNet101 model achieved comparable or superior performance to some reported results, despite not using transfer learning or fine-tuning, underscoring the potential for transfer learning to yield even more substantial improvements. It was also seen that MobileNetV2 AML transfer-trained model emerged BIC, when compared to some of the denser models evaluated, e.g. ResNet101 (42.5M parameters), Vision Transformer (85.6M parameters). This is in stark contrast with studies suggesting that deeper networks typically achieve higher accuracy.

## Data Availability

Complete dataset, model, and code are made available. The code for this research is available online at this GitHub link (https://github.com/AlphaKappaDelta81/TransferLearning), and the model weights can be found at the Zenodo repository (DOI: 10.5281/zenodo.15388608). The model weights are also stored in a Hugging Face repository (https://huggingface.co/AJ-MCL570s/TF_Learning_ALL_AML/tree/main) to ensure the reproducibility and validity of the results.

